# Population estimates of the number of adults in the United Kingdom with a hearing loss updated using 2021 and 2022 census data

**DOI:** 10.1101/2024.01.26.24301819

**Authors:** Michael A Akeroyd, Kevin J Munro

## Abstract

We update an earlier publication (Akeroyd MA et al., Int J Audiol. 2014 53(1):60-1) to derive new estimates of the number of adults in the UK with a hearing loss, using population data from the 2021/2022 UK censuses and prevalence data from the UK National Study of Hearing (Akeroyd MA et al. Trends In Hearing 2019 Jan-Dec;23:2331216519887614). Setting a criterion of a four-frequency hearing level >= 35 dB in the better ear gives an estimate of 4.6 million adults aged 18–80, which is a 17% increase on the earlier value from the 2011 census. With a criterion of >= 20 dB hearing level, the estimate is 12.3 million in total, or 1 in 4 of the population aged 18-80. The revised numbers highlight just how common hearing problems are in the population.

## MAIN TEXT

The release of data from the latest UK censuses, conducted in 2022 for Scotland and 2021 for the remainder of the UK, provides an opportunity to update our earlier publication on the population estimates of the number of adults with hearing loss (Akeroyd et al, 2013). As there, our method was to multiply the prevalence data from the National Study of Hearing (“NSH”; see tables B5124-12 to B5124-23 in Akeroyd et al., 2019) by the number of adults from the censuses. The NSH’s prevalence data are banded by decades of age. The England, Welsh and Northern Irish census data are reported year-by-year of age (Office for National Statistics, 2023; Northern Ireland Statistics & Research Agency, 2022), and so we summed those by NSH age band. The Scottish census, however, has so far only reported data in 5-year bands (Scotland’s Census, 2023), and those bands were defined slightly differently to the NSH’s (e.g. 30-34 and 35-39 years versus 31-40 years). To adjust for this difference, we fitted a cubic-spline function to the census data, to interpolate year-by-year values, then summed those by NSH age bands. For consistency with our earlier work we used a hearing loss of 35 dB or more in the better of the two ears (averaged over 500, 1000, 2000, 4000 Hz). For comparison we also recomputed the numbers from 2011 to cover the while United Kingdom (Office for National Statistics, 2013); though the NSH was not conducted in Northern Ireland, in the absence of evidence to the contrary we assumed its prevalence data were applicable there.

The results are set out in Table 1. The total estimate is 4.6 million adults (aged 18– 80) with a hearing level 35 dB or more in their better ear. This is a 17% increase on the earlier value from the 2011 census (Akeroyd et al., 2014), even though the total population aged 18-80 in this age band has only increased by 6%, from 47 million to 50 million. The percentage changes vary dramatically by age band, namely +21%, +7%, +28% for age bands of 50s, 60s, and 70s, respectively. The reason is that the changes by band directly reflect the ageing of two major peaks in UK birth rate, in 1947 and around 1964 (Office for National Statistics, 2015). As the population ages, these peaks enter then leave the age bands in the NSH calculations. The 51-60 band in 2021/22 now encompasses the 1960’s peak and the 71-80 band encompasses the 1947 peak, but the 61-70 band has lost the 1947 peak.

**Table 1.**
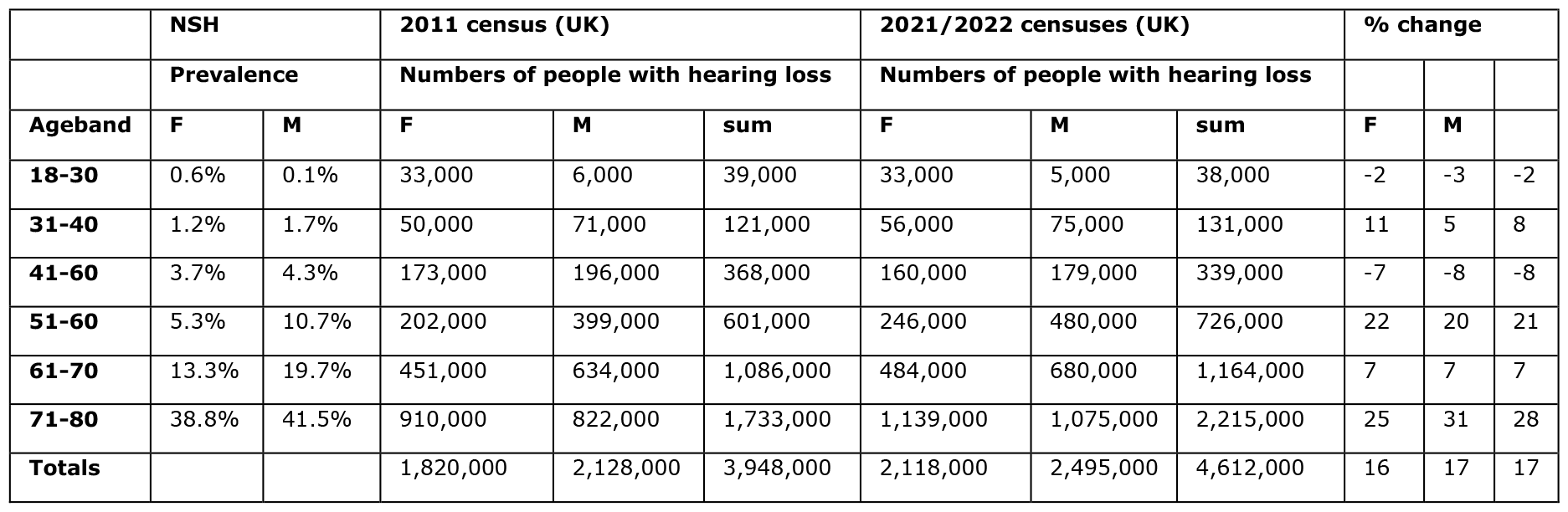
The expected number of adults aged 18–80 in the UK with hearing losses of at least 35 dB in the better ear, given the prevalence data in the NSH and the total population numbers from the 2011 census and the 2021/2022 census. The numbers are rounded to the nearest thousand. The three right-most columns report the percentage increase from 2011 to 2021/2022.

|  | NSH |  | 2011 census (UK) |  |  | 2021/2022 censuses (UK) |  |  | % change |  |  |
| --- | --- | --- | --- | --- | --- | --- | --- | --- | --- | --- | --- |
|  | Prevalence |  | Numbers of people with hearing loss |  |  | Numbers of people with hearing loss |  |  |  |  |  |
| Ageband | F | M | F | M | sum | F | M | sum | F | M |  |
| 18-30 | 0.6% | 0.1% | 33,000 | 6,000 | 39,000 | 33,000 | 5,000 | 38,000 | -2 | -3 | -2 |
| 31-40 | 1.2% | 1.7% | 50,000 | 71,000 | 121,000 | 56,000 | 75,000 | 131,000 | 11 | 5 | 8 |
| 41-60 | 3.7% | 4.3% | 173,000 | 196,000 | 368,000 | 160,000 | 179,000 | 339,000 | -7 | -8 | -8 |
| 51-60 | 5.3% | 10.7% | 202,000 | 399,000 | 601,000 | 246,000 | 480,000 | 726,000 | 22 | 20 | 21 |
| 61-70 | 13.3% | 19.7% | 451,000 | 634,000 | 1,086,000 | 484,000 | 680,000 | 1,164,000 | 7 | 7 | 7 |
| 71-80 | 38.8% | 41.5% | 910,000 | 822,000 | 1,733,000 | 1,139,000 | 1,075,000 | 2,215,000 | 25 | 31 | 28 |
| Totals |  |  | 1,820,000 | 2,128,000 | 3,948,000 | 2,118,000 | 2,495,000 | 4,612,000 | 16 | 17 | 17 |

In recent years there has been an increased interest in a criterion of 20 dB (better ear, across 500, 1000, 2000 and 4000 Hz) for calculating the prevalence of hearing loss, for example in the Global Burden of Disease (GBD Hearing Loss Collaborators, 2021). Figure 1 shows the numbers of people with a hearing loss according to this definition. They are 12.3 million in total, or 1 in 4 of the population aged 18-80. The numbers will be much greater still – 18 million, or 1 in 3 -- if those with 20 dB in the poorer ear are included (c.f. Supplementary Figure 1 for another example). Many such listeners will have hearing difficulties at listening in noise, sound localisation, etc. even if their better ear has a “normal” audiogram. This analysis highlights just how common numerically small, but still potentially significant to an individual, hearing losses actually are. Indeed, it is sometimes un-appreciated just how likely hearing losses are with ageing for a 20-dB criterion, especially for higher frequencies: for example, the NSH data indicates that it is near-certain (98%) that *everyone* aged 71-80 will have a hearing level of at least 20 dB in their better ear averaged across 4, 6 and 8 kHz.

**Figure 1.**
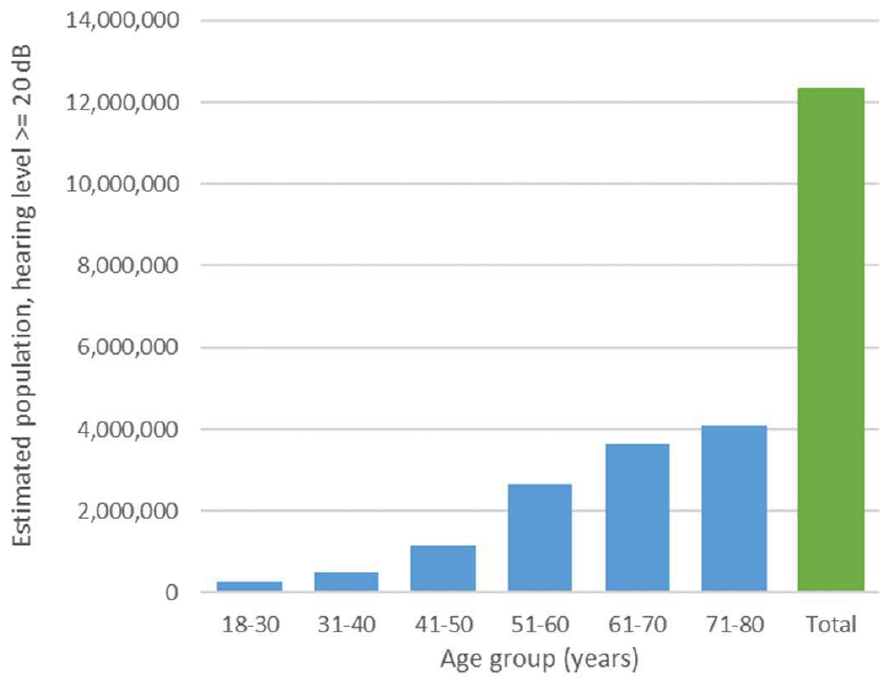
Expected number of adults (age 18-80) in the UK with hearing losses of at least 20 dB in the better ear, given the prevalence data in the NHS and the total population numbers from the 2021/2022 census. This does not include adults with asymmetric hearing losses who may experience hearing problems even if better hearing ear is within normal limits.

**SUPPLEMENT FIGURE 1.**
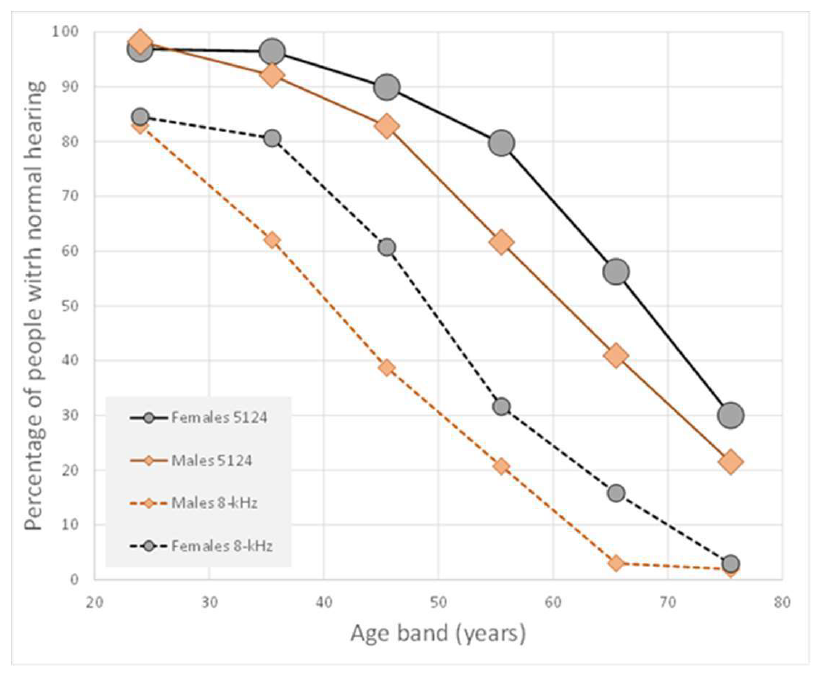
Percentage of males or females adults (aged 18-80) in England, Scotland and Wales with hearing level of at least 20 dB in the better ear (average of pure tone hearing threshold levels over 500, 1000, 2000 and 4000 Hz) as a function of age (the x-value is the midpoint of the age band). Also shown are data for pure tone hearing threshold level at 8000 Hz, indicating some degree of hearing loss, even if not amenable with current hearing aid technology. Data extracted from National study of hearing (Davis, 1995, Akeroyd et al. 2019)

In our earlier work we cautioned that all these values can only be estimates, as the prevalence data in the NSH were collected in the 1980s (Davis, 1995; Akeroyd et al, 2019). There are multiple reasons why the values may have changed either upwards or downwards – e.g. changes in general health, long-term changes in the economy away from noisy industry, noise-at-work regulations, lifestyle choices in leisure listening to loud music. Nevertheless, despite its age the NSH still remains the primary *audiological* data available in the UK, and therefore is the only data set that detailed calculations can be based on: the UK Biobank used a triple digit speech-in-noise test (Dawes et al., 2014) while the English Longitudinal Study of Ageing used a screening device set to 1-kHz and 3 kHz (Tsimpida et al 2020). But, there are both similarities and differences to the latest nationally-representative prevalence data in comparable countries. For instance, for the “CONSTANCES” study in France, which were collected between 2011 and 2019, the prevalence-by-age curve for males closely matches those that from the NSH, but for females, the French values are almost 10% greater than the NSH’s for 50’s and 60’s (Lisan et al., 2022). The resulting uncertainty as to which way the prevalence of hearing loss in the UK has actually changed since the 1980s emphasises the paucity of current epidemiological data (Whitty, 2023) and indicates that a suitable new UK national study of hearing needs be considered.

## Data Availability

All data produced in the present study are available upon reasonable request to the authors

https://doi.org/10.1177/2331216519887614

https://www.nisra.gov.uk/publications/census-2021-main-statistics-demography-tables-age-and-sex

https://www.ons.gov.uk/peoplepopulationandcommunity/populationandmigration/populationestimates/datasets/2011censuspopulationestimatesbysingleyearofageandsexforlocalauthoritiesintheunitedkingdom

https://www.ons.gov.uk/datasets/RM121/editions/2021/versions/1

https://www.scotlandscensus.gov.uk/2022-results/scotland-s-census-2022-rounded-population-estimates/

## ACKNOWLEDGEMENTS

Both authors contributed equally to the conceptualisation, analyses and writing. This research was supported by the National Institute for Health and Care Research (NIHR) Nottingham Biomedical Research Centre and Nottingham Biomedical Research Centre. The views expressed in this article are those of the authors and not necessarily those of the NHS, the NIHR, or the Department of Health and Social Care.

## DECLARATION OF INTEREST

None

## Notes

### Competing Interest Statement

The authors have declared no competing interest.

### Author Declarations

Data was obtained from recent UK censuses and the UKL National Study of Hearing https://www.nisra.gov.uk/publications/census-2021-main-statistics-demography-tables-age-and-sex https://www.ons.gov.uk/peoplepopulationandcommunity/populationandmigration/populationestimates/datasets/2011censuspopulationestimatesbysingleyearofageandsexforlocalauthoritiesintheunitedkingdom https://www.ons.gov.uk/peoplepopulationandcommunity/birthsdeathsandmarriages/livebirths/articles/trendsinbirthsanddeathsoverthelastcentury/2015-07-15 https://www.ons.gov.uk/datasets/RM121/editions/2021/versions/1 https://www.scotlandscensus.gov.uk/2022-results/scotland-s-census-2022-rounded-population-estimates/ https://doi.org/10.3109/14992027.2013.850539

## REFERENCES

Akeroyd, M. A., Browning, G. G., Davis, A. C., & Haggard, M. P. (2019). Hearing in Adults: A Digital Reprint of the Main Report From the MRC National Study of Hearing. Trends in hearing, 23, 2331216519887614. 10.1177/2331216519887614

Akeroyd, M. A., Foreman, K., & Holman, J. A. (2014). Estimates of the number of adults in England, Wales, and Scotland with a hearing loss. International Journal of Audiology, 53(1), 60–61. 10.3109/14992027.2013.850539

Davis A. C. (1995). Hearing in adults. London, England: Whurr.

GBD 2019 Hearing Loss Collaborators. (2021). Hearing loss prevalence and years lived with disability, 1990–j2019:j findings from the Global Burden of Disease Study 2019. Lancet Mar 13;397(10278):996–1009. doi: 10.1016/S0140-6736(21)00516-X.

Lisan Q, Goldberg M, Lahlou G, Ozguler A, Lemonnier S, Jouven X, Zins M, Empana JP. Prevalence of Hearing Loss and Hearing Aid Use Among Adults in France in the CONSTANCES Study. JAMA Netw Open. 2022 Jun 1;5(6):e2217633. doi: 10.1001/jamanetworkopen.2022.17633. Erratum in: JAMA Netw Open. 2022 Jul 1;5(7):e2225053. PMID: 35713903; PMCID: PMC9206187.

McCormack A, Edmondson-Jones M, Fortnum H, Dawes P, Middleton H, Munro KJ, Moore DR. The prevalence of tinnitus and the relationship with neuroticism in a middle-aged UK population. J Psychosom Res. 2014 Jan;76(1):56–60. doi: 10.1016/j.jpsychores.2013.08.018. Epub 2013 Aug 31. PMID: 24360142.

Northern Ireland Statistics & Research Agency (2022). Census 2021 main statistics demography tables – age and sex. https://www.nisra.gov.uk/publications/census-2021-main-statistics-demography-tables-age-and-sex(accessed 2024/1/17)

Office for National Statistics (2013). 2011 Census: Population estimates by single year of age and sex for local authorities in the United Kingdom. https://www.ons.gov.uk/peoplepopulationandcommunity/populationandmigration/populationestimates/datasets/2011censuspopulationestimatesbysingleyearofageandsexforlocalauthoritiesintheunitedkingdom(accessed 2024/1/17)

Office for National Statistics (2015). Trends in births and deaths over the last century. https://www.ons.gov.uk/peoplepopulationandcommunity/birthsdeathsandmarriages/livebirths/articles/trendsinbirthsanddeathsoverthelastcentury/2015-07-15(accessed 2024/1/16)

Office for National Statistics (2023). Sex by age. https://www.ons.gov.uk/datasets/RM121/editions/2021/versions/1(accessed 2024/1/16)

Scotland’s Census (2023). Scotland’s Census 2022 - Rounded population estimates. https://www.scotlandscensus.gov.uk/2022-results/scotland-s-census-2022-rounded-population-estimates/(accessed 2024/1/16)

Tsimpida D, Kontopantelis E, Ashcroft D, Panagioti M. Comparison of Self-reported Measures of Hearing With an Objective Audiometric Measure in Adults in the English Longitudinal Study of Ageing. JAMA Netw Open. 2020 Aug 3;3(8):e2015009. doi: 10.1001/jamanetworkopen.2020.15009. PMID: 32852555; PMCID: PMC7453309.

Whitty, C. J. M. (2023). Chief Medical Officer’s Annual Report 2023: Health in an Ageing Society. London: Department of Health and Social Care (2023) http://www.gov.uk/government/publications/chief-medical-officers-annual-report-2023-health-in-an-ageing-society

